# Exploratory brain-wide analyses predicting binge drinking: A machine learning and targeted causal inference pipeline in two adolescent cohorts

**DOI:** 10.64898/2026.10.18.26365855

**Authors:** Rachel Visontay, Hollie R. Byrne, Emma K. Devine, Matthew Sunderland, Lei Fan, Wei Wen, Maria I. Perica, Susan Tapert, Duncan B. Clark, Kate Nooner, Kamran Afzali, Lindsay M. Squeglia, Louise Mewton

**Affiliations:** The Matilda Centre for Research in Mental Health and Substance Use, The University of Sydney. Sydney, Australia; Centre for Healthy Brain Ageing, UNSW; Department of Psychiatry and Behavioral Sciences, Medical University of South Carolina, Charleston, USA; University of California, San Diego, USA; Department of Psychiatry, University of Pittsburgh, Pittsburgh, PA; Department of Psychology, University of North Carolina Wilmington, Wilmington, NC, 28403, United States; The Research Institute of the McGill University Health Centre Montreal, Canada

**Author notes:** Correspondence to: Dr Rachel Visontay, The Matilda Centre for Research in Mental Health and Substance Use, The University of Sydney, Sydney, 2008, Australia. Joint senior authors (Lindsay M. Squeglia, Louise Mewton).

**Keywords:** neurobiology, MRI, binge drinking, adolescence, machine learning, causal inference

## Abstract

**Background:** A sizeable literature has found that neuroanatomical differences predict alcohol use outcomes in adolescence. However, due possibly to small samples and inconsistent/limited methodologies, it is still unclear which specific morphometric features are most predictive and if they are also *causally* implicated in binge drinking onset.

**Methods:** To differentially predict abstinence from binge drinking by age 16-17, a SuperLearner ensemble algorithm was fit to structural and diffusion MRI features at age 9-10 in the Adolescent Brain and Cognitive Development cohort (ABCD). A separate model was fit using the National Consortium on Alcohol and NeuroDevelopment in Adolescence (NCANDA), in which the MRI predictors were measured later (ages 12-17) but the developmental outcome (binge drinking by age 18) remained similar. The top ten predictors were extracted from each model, and the causal role of these features scrutinised using targeted maximum likelihood estimation (TMLE).

**Results:** Predictive performance in ABCD was poor (AUC=.585) but was stronger in NCANDA (AUC=.695). There was little overlap in predictive feature rankings between the cohorts. TMLE did not reveal a causal effect of the top ten morphological predictors on binge drinking in either cohort.

**Conclusions:** No evidence was found for causal contributions of any discrete macrostructural/microstructural brain feature in late childhood or adolescence to binge drinking. Using more sophisticated metrics of brain structure and function may yield better insights. Adding to previous evidence from youth cohorts, neuroanatomy may have greater predictive value for identifying those at risk of adolescent binge drinking when measured in adolescence, rather than late childhood.

## INTRODUCTION

Adolescence is a key period for the initiation and escalation of alcohol use, with early initiation being one of the strongest predictors of future alcohol use disorder (AUD). Early adolescent alcohol use is often characterised by less frequent, episodic binges, with 15-20% of adolescents experiencing drunkenness by age 15-16 (1,2). Binge drinking is associated with substantial harms, including alcohol-related blackouts, risky sexual behaviours, injuries, suicide, mental illness, as well as AUD and other drug disorders (3).

While numerous factors likely predispose adolescents to the initiation of binge drinking, dominant theories of adolescent alcohol use focus on neurobiological factors and maturational processes occurring in the adolescent brain. Adolescent neurodevelopment is characterised by earlier maturation of subcortical limbic regions contributing to reward and socioemotional systems, while prefrontal systems that modulate cognitive control tend to develop later. The imbalance in the development of these systems is hypothesised to create a propensity towards risk-taking behaviours, which are adaptive and necessary for independence, but also associated with health risk behaviours that may cause harm. These theories of adolescent risk-taking have been formalised in the dual systems (4) and maturational imbalance (5) hypotheses, which dominate brain-based theories of adolescent alcohol use.

Consistent with these theories, our recent systematic review of longitudinal structural and diffusion-weighted neuroimaging studies found that aberrant development of subcortical and frontal systems appeared to prospectively predict increased risk of early and escalating alcohol use in adolescence (6), these regions being associated with increased reward-seeking, risk-taking, and poorer decision-making. However, the specific neuroanatomical features implicated in the risk for adolescent alcohol use remain to be identified (7), with inconsistent findings and conflicting directions of effect currently observed in the literature (6). These inconsistencies may be due to relatively small sample sizes in prior research, as well as some evidence that alcohol use behaviours may be differentially mediated by reward and inhibitory control systems depending on whether they index more normative or problematic alcohol involvement (e.g., (8)). Additionally, existing studies vary in their use of brain volume, cortical thickness, and surface area measurements to index grey matter development, which may obscure important effects given the different developmental trajectories of each (9). Ideally, any study focused on neurobiological predictors of alcohol use would incorporate all three measures, as well as indicators of diffusion MRI-derived white matter integrity, also implicated in the development of adolescent alcohol use (7).

Traditional models struggle to accommodate such large sets of predictors due to threats from multicollinearity, overfitting, and the difficulty of correct specification of both interaction terms and functional form between each predictor and outcome. In contrast, machine learning approaches are ideally suited for use with high dimensional data. Cross-validation (repeatedly evaluating model fit in a separate portion of the sample to that used for model training) mitigates overfitting, and base algorithms can be employed that are robust to multicollinearity and that do not rely on explicit specification of functional forms and interaction terms.

Given the advantages of machine learning approaches, these have increasingly been applied in studies seeking to understand the neurobiological basis of adolescent alcohol use (10–12). Many of these studies have been based on the Adolescent Brain Cognitive Development Study (ABCD), but overall prediction accuracy of these brain-based machine learning models of adolescent alcohol initiation have tended to be poor. This may be due to a focus on earlier alcohol use milestones, such as alcohol sipping and first drink. As the ABCD cohort ages, there is now an opportunity to examine heavier drinking phenotypes, particularly binge drinking, which may be more related to neuroanatomical precursors, as indicated in samples of older adolescents (11).

While poor prediction may be a result of the alcohol use outcome in these studies, it may also be due to the age at which the brain-based predictors are collected. In ABCD, baseline neuroimaging assessment occurs at ages 9-10 years, which has the advantage of occurring before most adolescents have initiated alcohol use. However, this is also prior to the age at which neurodevelopment of frontal and limbic systems is theorised to diverge. For this reason, we also fit a separate algorithm using the National Consortium on Alcohol and NeuroDevelopment in Adolescence (NCANDA), a precursor to ABCD designed specifically to enhance understanding of alcohol use in adolescence and its relationship with the brain (13). From this cohort we analyse individuals in which the MRI predictors were measured later than in ABCD (ages 12-17) but the developmental outcome (binge drinking by age 18) remained similar.

Although the overall prediction of binge drinking offered by a brain-wide assembly of neuroanatomical features may not be strong, these models can be useful in identifying *which* morphometric features are most predictive of adolescent alcohol use. Consistent with major brain-based theories of adolescent alcohol use, prior studies have found that certain prefrontal and limbic features are associated with later alcohol initiation (11,14).

Understanding which brain-based features predispose individuals to binge drinking is crucial to ongoing efforts to improve identification of at-risk adolescents. But for developing worthwhile preventative efforts, we must know if these features are also *causally* implicated in the development of binge drinking – something traditional machine learning approaches cannot elucidate (15). Confounding is one of the biggest obstacles to being able to attribute a relationship’s observed functional form and strength directly to the effect of exposure on outcome (i.e., causal inference). To promote causal inference when using observational data, modern statistical techniques have been developed that mimic the randomisation mechanism of randomised controlled trials, mitigating the impact of confounders. However, these approaches have rarely been applied when examining neurodevelopmental–behaviour relationships in observational data (16).

In the current study we applied a consistent analytic pipeline to two developmentally distinct cohorts to examine the extent to which an ensemble machine learning approach can distinguish between adolescents who remain abstinent and those who engage in early initiation of binge drinking (excluding non-binge drinking youth who are using alcohol at lower levels), as well as the neuroanatomical features most important in the prediction of binge drinking. Following the identification of the most predictive features, we examined whether these features were causally implicated in binge drinking using targeted maximum likelihood estimation (TMLE).

## METHODS

### Study Design and Participants

ABCD is a multi-site (N=21) longitudinal study following a large adolescent cohort (N=11,860) in the U.S. Children were recruited when aged 9-11, from 2016-2018. For the current study, the ABCD 7.0 release was used, which contains near-complete data up to the eighth assessment wave (N=8,564; age 16-18). Parents or caregivers provided informed consent and all participants gave assent. Those with structural and diffusion MRI data at baseline that passed ABCD imaging quality control inclusion criteria were included in ComBat neuroimaging data harmonisation to remove site effects.

After harmonisation, those who had consumed a full drink at the baseline wave, and those for whom it was not possible to ascertain engagement in binge drinking by the eighth wave, were further excluded. Finally, those who had consumed a full drink by the eighth wave but not binged (defined here and in NCANDA as at least one occasion of >=4 drinks for females; >=5 drinks for males), were removed, such that the model could attempt to distinguish between binge drinkers and abstainers while excluding intermediate alcohol users. For subsequent TMLE analyses, random removal of siblings was performed (to satisfy the independence assumption underlying TMLE’s variance estimation) as was removal of those with missing covariate data.

The older adolescent cohort used was NCANDA, a cohort designed to study adolescent development and alcohol involvement across 5 sites, with enrichment for youth at varying levels of alcohol use risk. Here too, those without valid MRI data were excluded and ComBat harmonisation was performed (excluding diffusion data, which was already provided in harmonised form).

The rationale behind selecting the NCANDA study was to perform analyses in a cohort in which the brain was scanned later (ages 12-17) than in ABCD, but where the developmental outcome remained binge drinking before adulthood. To accommodate this goal, those who were already aged 18 or above at baseline were removed from the initial sample of 831 individuals, leaving just those for whom scans were obtained at age 12-17. Those who had consumed a full drink at baseline and those who were not confirmed abstainers or binge drinkers by the first wave after turning 18 were also removed. See Figures 1a and 1b for flow diagrams of participant exclusion.

**Figure 1a.**
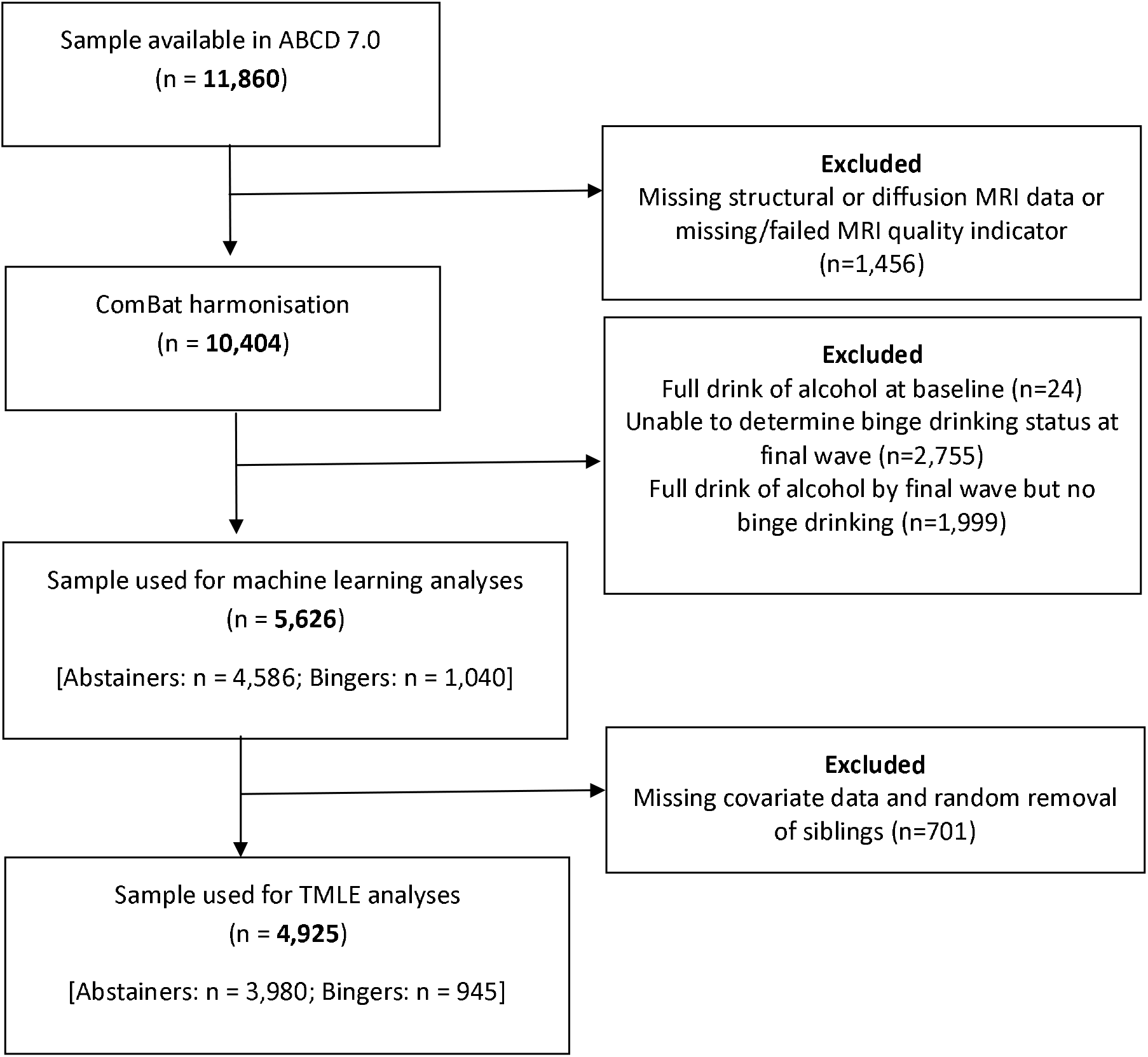
Eligibility flow chart for analyses in ABCD

**Figure 1b.**
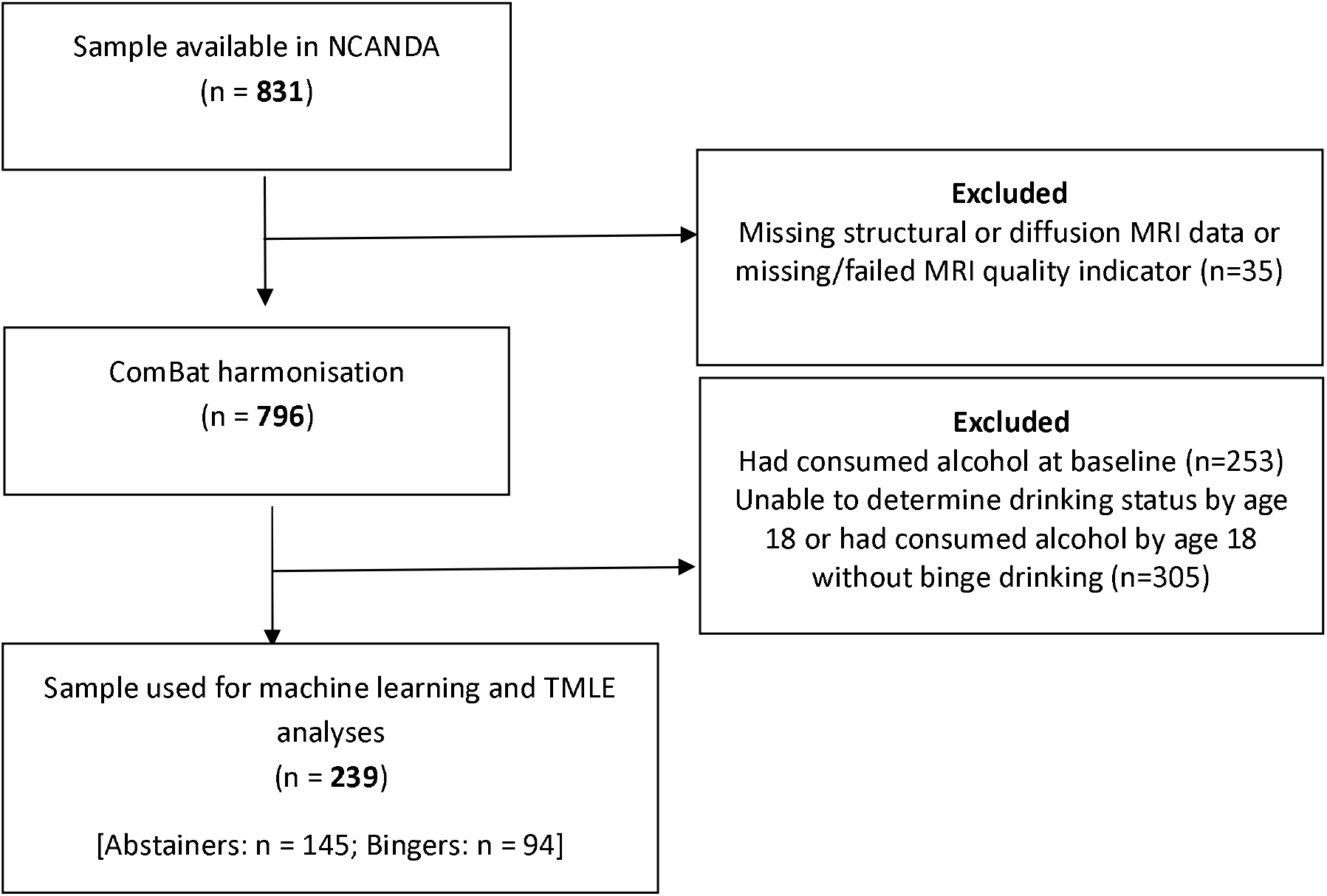
Eligibility flow chart for analyses in NCANDA

### Measures

#### MRI-derived Predictors (Machine learning and TMLE)

An exploratory, brain-wide set of predictors was derived from structural and diffusion MRI data acquired at study baseline for ABCD. The processing of this data, described in detail elsewhere (17), involved cortical parcellation and subcortical segmentation of T1-weighted structural imaging data using FreeSurfer (v7.1.1). This resulted in 218 cortical grey matter measures (cortical volume, surface area, and thickness measurements mapped to 68 cortical regions in the Desikan-Killiany atlas (18), as well as subcortical volumes for 14 regions derived from FreeSurfer’s automated subcortical segmentation). Additionally, 93 diffusion tensor imaging (DTI) derived variables were included, based on 14 bilateral white matter tracts and 3 unilateral white matter tracts defined using AtlasTrack (19), each indexed by fractional anisotropy (FA), mean diffusivity (MD), and radial diffusivity (RD). The total number of features was 311.

For NCANDA, 218 structural MRI features were included using the same FreeSurfer processing and feature extraction methodology described for ABCD. DTI-derived variables included 81 measures in total, based on 27 white matter tracts from the JHU DTI white matter atlas (20), indexed by FA, MD, and RD. The total number of features was 299.

#### Outcomes (Machine learning and TMLE)

The binge-drinking outcome (as described above) was determined using the last wave of data in ABCD (at which most participants were aged 16-17), and the first wave at which participants were 18 in NCANDA. Abstinence was defined as not having consumed a full drink by this same age.

#### Covariates (ComBat Harmonisation and TMLE)

Covariates of interest for the TMLE analyses in ABCD were: sex (binary), race/ethnicity (five categories), highest parental education level across caregivers (five levels), whether the child’s religion has prohibitions on alcohol (binary), whether the child had ever sipped alcohol at baseline (binary), standardised intracranial volume, and standardised age. Missing data in these covariates were converted to a ‘missing’ indicator level (there was no missing data in the age variable). For NCANDA, covariates differed slightly due to data availability: sex (binary), race (four categories), Hispanic ethnicity (binary), highest parental education level (five levels), standardised intracranial volume, and standardised age.

### Statistical Analyses

#### Machine Learning

An overview of the analysis pipeline is presented in Figure 2. First, MRI data were harmonised to account for batch effects using ComBat (21) (see Supplement for details). Next, all predictors were standardised. A SuperLearner algorithm (composed of XGBoost, Random Forests, kernel Support Vector Machine, and elastic-net base learners) was used to predict binge drinking using the full set of baseline morphometric predictors. This generated a final ‘ensemble’ model (i.e., a model that combines the base learners in a weighted fashion based on their individual performance, rather than selecting the single best performer – see details in Figure 2 and Supplement).

**Figure 2.**
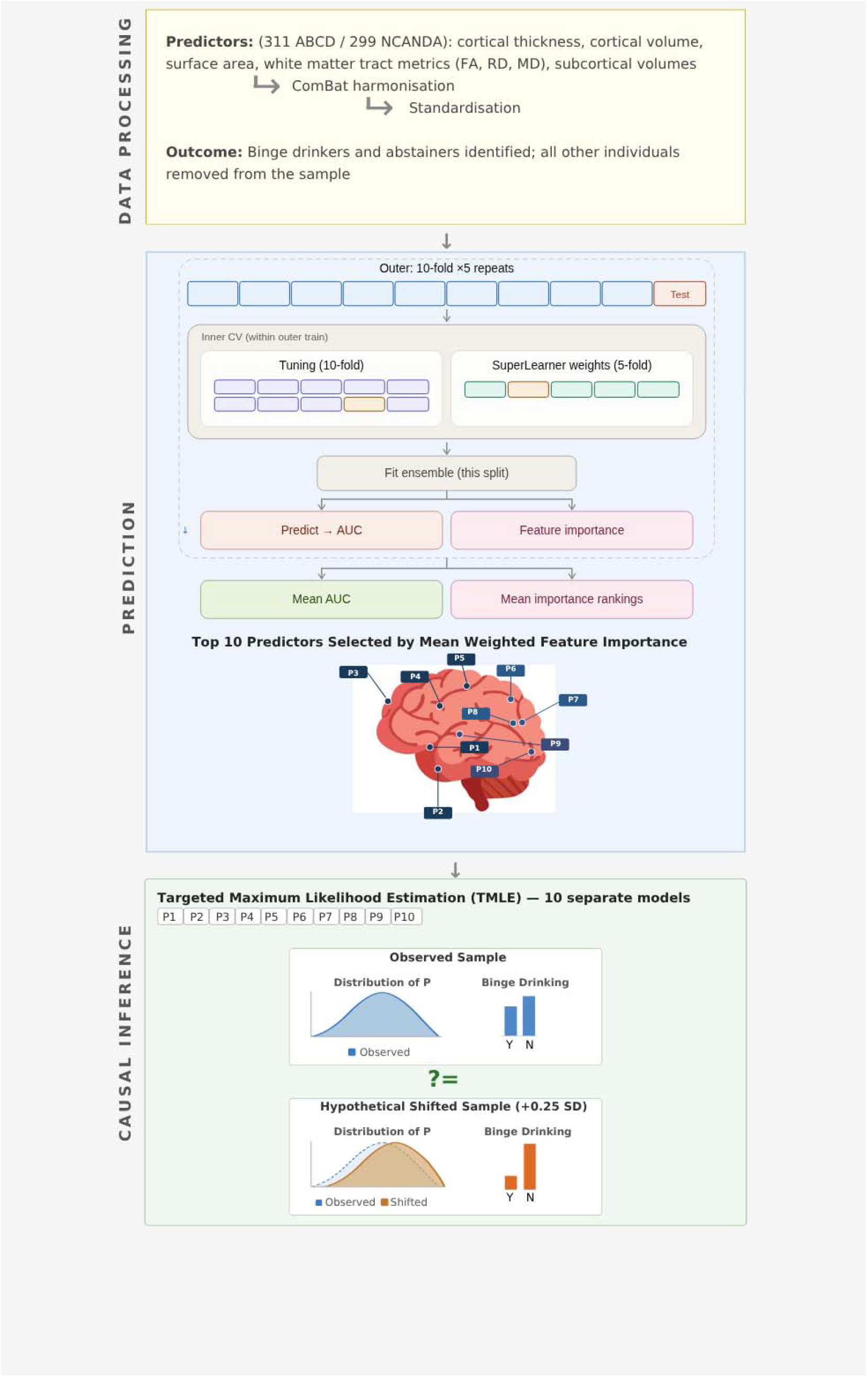
Machine learning and causal inference pipeline

Area Under the Curve (AUC) metrics were generated for the best ensemble model in the testing set. To determine the importance of brain features associated with binge drinking, the importance of each feature for each base learner was calculated within each outer fold using base learner-specific importance metrics (see Supplement for details). These metrics were then normalised to be comparable across base learners, before being weighted by the weights assigned to each base learner by the SuperLearner, producing a single mean weighted importance metric and associated standard deviation for each predictor (22).

#### Targeted Maximum Likelihood Estimation

From these resulting ranked lists of feature importance, the top predictors were extracted for further examination of a potentially causal role. In the absence of a clear gap or discontinuity in importance between subsequent predictors on the ranked list, the top ten features were chosen as a pragmatic cutoff. TMLE is considered a doubly robust method for causal inference (see Supplement), which means that only one of either the exposure or outcome models need be consistently estimated for the final estimate to be unbiased (23).

Because TMLE is non-parametric, and because it also has a strong positivity assumption (i.e., it can only draw conclusions about exposure levels actually represented in the data), it does not lend itself to estimates of the form ‘a 1-unit increase in the exposure is associated with a Y-unit increase in the outcome’. Instead, in the context of a continuous exposure (e.g., a given morphometric feature), we generally estimate the effect of a *shift in the distribution* of the exposure (see bottom panel of Figure 2) (24,25).

In the case of a categorical outcome, TMLE estimates what effect manipulating the exposure via some kind of hypothetical population intervention would have on the proportion of the sample developing the outcome (‘risk for outcome’). In this case, that would mean evaluating whether there is a significant difference between the observed proportion of bingers in the actual sample and the estimated proportion of bingers in a world in which everyone had brain feature A shifted up by a certain value δ. If brain morphometry is a causal factor, then population interventions on early-life exposures that affect their development may be an important preventative for harmful binge drinking. We chose a 0.25 standard deviation distributional increase in exposure as the hypothetical shift of interest, corresponding to a very strong hypothetical intervention on a given baseline morphometric feature in the population (comparable to the effect size of very preterm birth on baseline neuroanatomy in ABCD (26)). While it is unlikely that any intervention would have such a strong effect on a given morphometric feature, failure of such a hypothetical intervention to produce predicted changes in binge drinking numbers in the sample indicates that the relationship between that brain feature and adolescent drinking is unlikely to be causal.

Analyses were conducted in R version 4.4.2 using packages neuroCombat (21), SuperLearner (27), and lmtp (28), and brain plots created using packages ggseg (29) and ggAtlasTrack (30).

## RESULTS

### Machine learning

Tables 1 and 2 detail the participant characteristics for ABCD and NCANDA respectively.

**Table 1.** Participant characteristics of ABCD sample included in machine learning analyses.

| Characteristic | Group |  |  |
| --- | --- | --- | --- |
|  | Total<br>N = 5626<sup>1</sup> | Abstainers<br>N = 4586<sup>1</sup> | Binge drinkers<br>N = 1040<sup>1</sup> |
| **Sex** |  |  |  |
| Male | 2,940 (52%) | 2,455 (54%) | 485 (47%) |
| Female | 2,686 (48%) | 2,131 (46%) | 555 (53%) |
| **Age** |  |  |  |
| Mean (SD) | 9.9 (0.6) | 9.9 (0.6) | 10.2 (0.6) |
| **Ethnicity<sup>2</sup>** |  |  |  |
| Hispanic | 1,091 (19%) | 897 (20%) | 194 (19%) |
| White | 3,096 (55%) | 2,410 (53%) | 686 (66%) |
| Black | 796 (14%) | 751 (16%) | 45 (4%) |
| Other | 643 (11%) | 528 (12%) | 115 (11%) |
| **Parental education** |  |  |  |
| Up to high school (No diploma) | 239 (4%) | 208 (5%) | 31 (3%) |
| High school/GED | 452 (8.0%) | 393 (9%) | 59 (6%) |
| Some college | 1,320 (23%) | 1,102 (24%) | 218 (21%) |
| Bachelor's degree | 1,562 (28%) | 1,288 (28%) | 274 (26%) |
| Graduate school/Professional degree | 2,043 (36%) | 1,585 (35%) | 458 (44%) |
| Missing | 10 (0.2%) | 10 (0.2%) | 0 (0%) |
| **Religious alcohol prohibition** |  |  |  |
| No | 3,423 (61%) | 2,636 (57%) | 787 (76%) |
| Yes | 1,557 (28%) | 1,413 (31%) | 144 (14%) |
| Don't know/Missing | 646 (11%) | 537 (12%) | 109 (10%) |
| **Baseline alcohol sipping** |  |  |  |
| No | 4,269 (76%) | 3,645 (79%) | 624 (60%) |
| Yes | 1,157 (21%) | 749 (16%) | 408 (39%) |
| Missing | 200 (3.6%) | 192 (4.2%) | 8 (0.8%) |

<sup>1</sup>n (%)
<sup>2</sup>Categories collapsed due to low cell counts

**Table 2.** Participant characteristics of NCANDA sample included in machine learning analyses.

| Characteristic | Group |  |  |
| --- | --- | --- | --- |
|  | Total<br>N = 239<sup>1</sup> | Abstainers<br>N = 145<sup>1</sup> | Binge<br>drinkers<br>N = 94<sup>1</sup> |
| **Sex** |  |  |  |
| Male | 119<br>(50%) | 67 (46%) | 52 (55%) |
| Female | 120<br>(50%) | 78 (54%) | 42 (45%) |
| **Age** |  |  |  |
| Mean (SD) | 14.4 (1.4) | 14.5 (1.5) | 14.2 (1.1) |
| **Race<sup>2</sup>** |  |  |  |
| White | 166<br>(69%) | 91 (63%) | 75 (80%) |
| Other | 73 (31%) | 54 (37%) | 19 (20%) |
| **Hispanic ethnicity** |  |  |  |
| No | 211<br>(88%) | 128 (88%) | 83 (88%) |
| Yes | 28 (12%) | 17 (12%) | 11 (12%) |
| **Parental education<sup>2</sup>** |  |  |  |
| Less than high school, High school/GED, or Associate degree | 56 (23%) | 40 (28%) | 16 (17%) |
| Bachelor's degree (B.A., B.S.) | 95 (40%) | 53 (37%) | 42 (45%) |
| Master's degree, Doctorate, Professional degree, or 'Other' | 88 (37%) | 52 (36%) | 36 (38%) |

<sup>1</sup>n (%)<sup>2</sup>Categories collapsed due to low cell counts

The cross-validated SuperLearner in ABCD achieved an AUC of 0.585, indicating prediction just above chance. The top ten importance rankings for each morphometric feature in ABCD – based on weighted ranked feature importance aggregated across algorithms – are presented in Table 3 (see Supplemental spreadsheet 1 for full list), with the full set of predictors visually depicted in Figure 3a in which darker cortical, subcortical, or white matter features reflect greater predictive importance. These importance scores reflect magnitude of importance, but do not incorporate direction of effect.

**Figure 3a.**
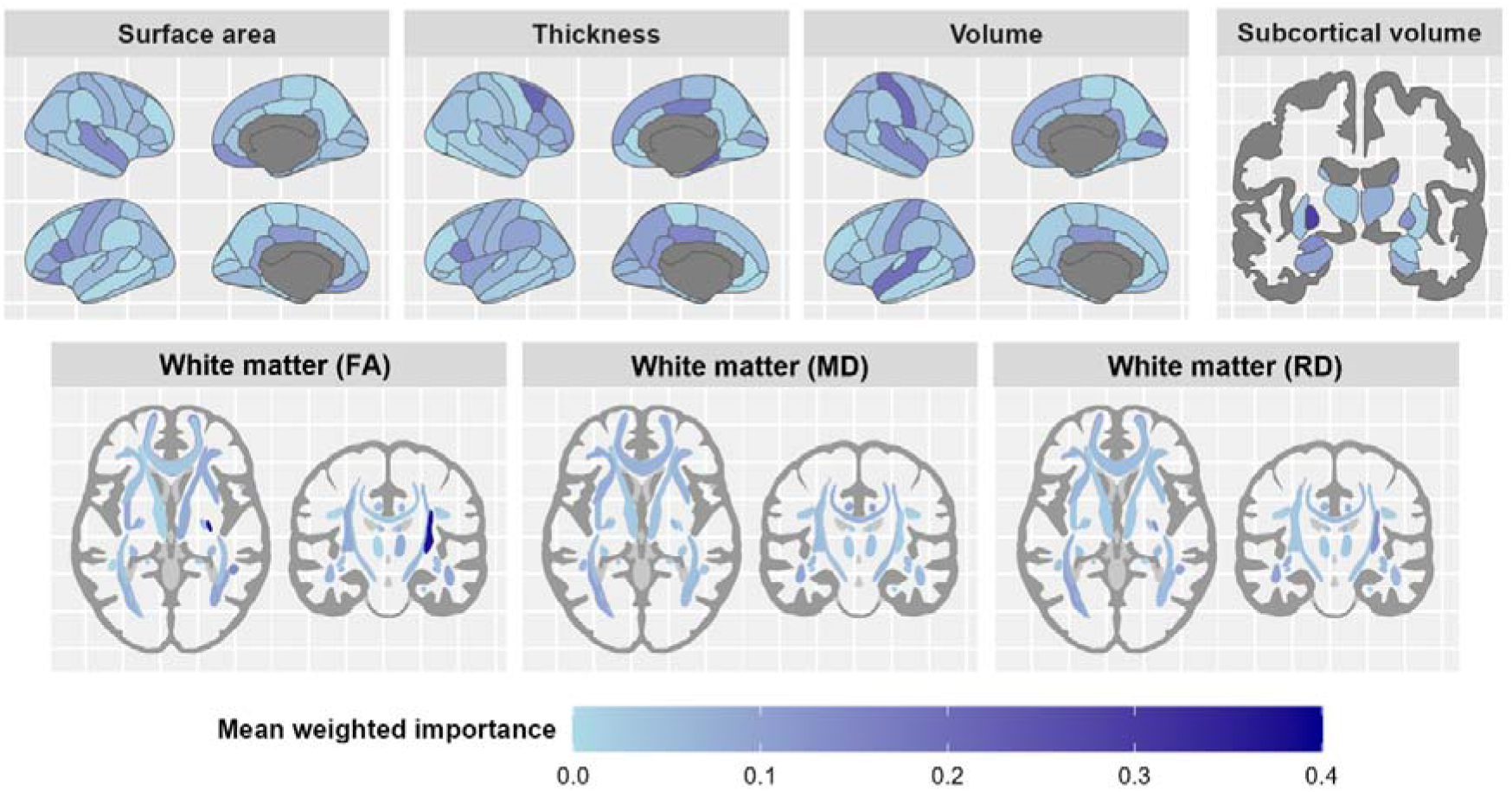
Feature importance in ABCD SuperLearner

**Table 3.** Estimated proportion of binge drinkers in ABCD corresponding to a .25SD increase in morphometric feature (observed proportion is .192)

| **Morphometric feature** | **Estimate** | **95% CI** |
| --- | --- | --- |
| DTI Fractional Anisotropy - Superior Corticostriate (right) | 0.189 | 0.158, 0.219 |
| Subcortical Volume - Pallidum (left) | 0.198 | 0.166, 0.229 |
| Cortical Thickness - Parahippocampus (right) | 0.192 | 0.162, 0.222 |
| Cortical Thickness - Caudal middle frontal lobe (right) | 0.188 | 0.158, 0.217 |
| Cortical Volume - Precuneus (right) | 0.193 | 0.162, 0.224 |
| Cortical Volume - Superior temporal lobe (left) | 0.186 | 0.154, 0.218 |
| DTI Fractional Anisotropy - Uncinate Fasciculus (right) | 0.189 | 0.156, 0.221 |
| Cortical Thickness - Postcentral gyrus (right) | 0.191 | 0.16, 0.223 |
| Cortical Surface Area - Pars opercularis (left) | 0.195 | 0.165, 0.225 |
| Cortical Thickness - Pars opercularis (left) | 0.189 | 0.156, 0.222 |

In general, more informative predictors in ABCD were dispersed across different metrices and were located diffusely across the brain. The strongest signals came from the superior corticostriate white matter tract (as indexed by FA) and volume of the left pallidum.

For NCANDA, the AUC was 0.695. The top ten importance rankings for each morphometric feature in NCANDA – based on weighted ranked feature importance aggregated across algorithms – are presented in Table 4 (see Supplemental spreadsheet 2 for full list), with the full set of predictors visually depicted in Figure 3b (importance scores are not on the same scale across datasets and are not comparable). Again, the most important cortical predictors were dispersed across lobes. Of note, the left isthmus cingulate had a clear predictive signal across volume, thickness, and surface area metrics. Subcortical volumes featured more strongly amongst the top predictors in NCANDA compared to ABCD (in particular, there was a strong bilateral signal from the putamen). Conversely, white matter featured less prominently in NCANDA, with the highest predictive values belong to tracts linking predictive subcortical regions, including the genu of the corpus callosum, the anterior limb of the internal capsule, and the posterior thalamic radiation.

**Figure 3b.**
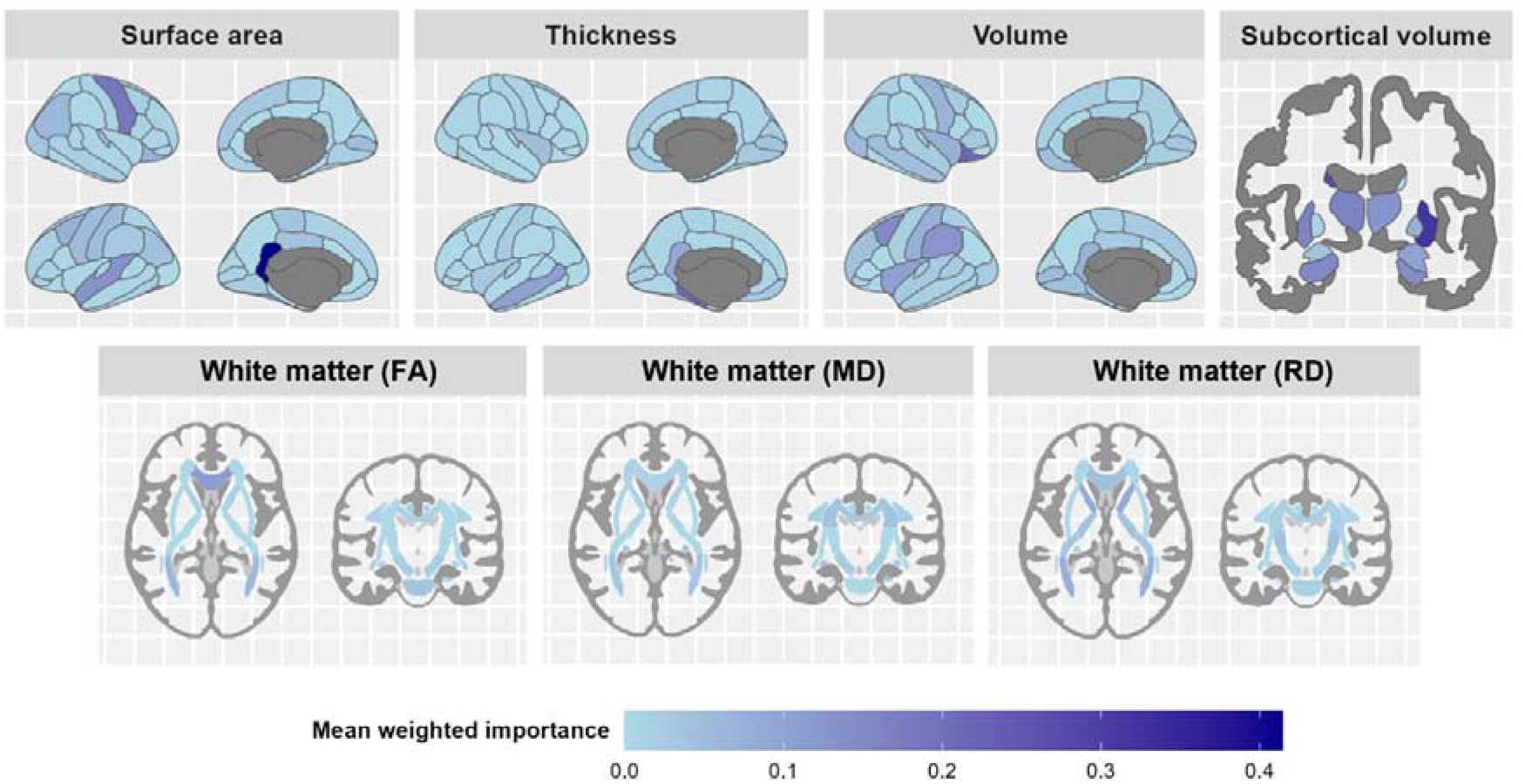
Feature importance in NCANDA SuperLearner

**Table 4.** Estimated proportion of binge drinkers in NCANDA corresponding to a .25SD increase in morphometric feature (observed proportion is .393)

| **Morphometric feature** | **Estimate** | **95% CI** |
| --- | --- | --- |
| Cortical Surface Area - Isthmus cingulate cortex (left) | 0.428 | 0.312, 0.512 |
| Subcortical Volume - Putamen (right) | 0.421 | 0.304, 0.517 |
| Subcortical Volume - Caudate (left) | 0.412 | 0.295, 0.527 |
| Cortical Volume - Lateral orbito-frontal lobe (right) | 0.439 | 0.312, 0.533 |
| Cortical Thickness - Parahippocampus (left) | 0.431 | 0.337, 0.517 |
| Cortical Surface Area - Precentral gyrus (right) | 0.418 | 0.300, 0.529 |
| Subcortical Volume - Thalamus (left) | 0.410 | 0.305, 0.513 |
| Subcortical Volume - Putamen (left) | 0.425 | 0.310, 0.526 |
| Subcortical Volume - Hippocampus (left) | 0.418 | 0.309, 0.513 |
| Cortical Volume - Caudal middle frontal lobe (left) | 0.416 | 0.300, 0.524 |

There were no features that appeared in the top ten predictors for both ABCD and NCANDA.

### Causal analysis of top predictors

The observed mean risk in ABCD was .192, corresponding simply to the ratio of bingers to abstainers in the final sample. The effect on the risk of binge drinking produced by a .25 SD increase was estimated for each of the top ten predictive features independently. Even under this large hypothetical effect size, manipulating the exposure did not produce a significant shift in the proportion of the sample predicted to engage in binge drinking (i.e., the confidence interval for the estimated risk still included the observed risk; Table 3). For example, a .25 SD increase across the distribution of DTI (FA) of the superior corticostriate (right) was estimated to reduce proportion of binge drinkers in the sample by just .003.

For NCANDA, the observed mean risk was .393. A .25 SD increase across the exposure distribution also did not lead to any significant shifts in predicted binge drinking across the sample for any of the top ten predictive features (although all estimates were consistent with an increase across the distribution of the morphometric feature leading to increased cases of binge drinking; Table 4).

## DISCUSSION

In two large, developmentally informative cohorts from the general population, a rigorous pipeline combining machine learning and causal inference indicated poor predictive utility and causal contributions of earlier brain morphometry for the development of adolescent binge drinking. The combined brain-wide set of predictors distinguished between abstainers and binge drinkers at levels just above chance in the younger ABCD cohort, but had fair performance in the NCANDA sample, for whom MRI scans indexed early-to-mid-adolescent neurodevelopment. There was little overlap between the most important predictive features in the two cohorts, although some identified variables of importance were consistent with the existing literature, as well as major theories of adolescent alcohol use. However, subsequent TMLE analyses did not provide evidence of causal contributions of any discrete macrostructural or microstructural feature in late childhood or early adolescence to adolescent binge drinking.

The poor predictive utility of baseline brain morphometry in ABCD aligns with previous studies in this cohort that have examined alcohol and other substance use outcomes (31,32), as well as research in other similarly-aged cohorts (33). In NCANDA, however, predictive utility of brain morphometry was stronger, aligning with machine learning research in other slightly older cohorts such as the IMAGEN dataset (11). It may be that neuroanatomy measured in early adolescence is more informative for identifying those at risk of teen binge drinking than when measured in late childhood. Additionally, white matter regions of interested tended to rank more highly in predictive importance in ABCD than in NCANDA, while the opposite was true of subcortical regions.

In general, the list of top predictive features in both cohorts were diffusely distributed across the brain, with mixed findings across different metrics for cortical and white matter regions of interest. This is in keeping with other machine learning studies, such as the work that has been carried out in the IMAGEN cohort finding that important predictors for adolescent alcohol misuse are diffuse and heterogenous (11).

The lack of clear patterns was particularly true of ABCD, in keeping with the poor overall predictive performance of the algorithm. That said, there was a relatively strong predictive signal from FA in the superior corticostriate tract, implicated in self-regulation (34), as well as from pallidal volume. This latter finding aligns with previous research in ABCD finding the pallidum to be associated with substance use and sensation seeking (35). In other work, FA in white matter proximal to the pallidum has been linked to the initiation of binge drinking in adolescents (36).

In keeping with its stronger predictive performance, several of the most important features in NCANDA were consistent with what would be expected based on the literature. For example, volume of the right lateral orbito-frontal lobe was the fourth most important predictor, which has been associated, albeit inconsistently, with adolescent alcohol use (7), with some work positing it may be the ratio between orbitofrontal and amygdala volumes specifically implicated in adolescent drinking (37). The left isthmus cingulate – involved in memory and emotion processing – was also dominant in the top NCANDA predictors, featuring across surface area, volume, and thickness. Volumes of the left and right putamen were also important – a region involved in habit formation and reward processing (38).

Importantly though, we found no evidence that these features are *causally* implicated in the development of binge drinking. Even with a very large hypothetical intervention on morphometric features in the population, there was almost no perceptible shift in predicted binge drinking numbers across either sample. This is consistent with recent studies that have found social and familial factors to be much more relevant in the development of substance use in this age group (31,32).

At face value, the lack of causal neurobiological mechanisms underpinning adolescent alcohol use is at odds with neurodevelopmental ‘imbalance’ theories of substance use. However, it may be that discrete atlas-based macrostructural or microstructural features are too coarse to capture the complex, distributed processes giving rise to alcohol use behaviours. Employing network-based metrics of brain structure and function that recognise the interconnected nature of the brain may yield better insights (e.g., (39,40)).

### Strengths

This study employed a robust approach to machine learning, isolating the focus to neuroanatomical predictors (instead of situating them amongst social and familial factors that may obscure signals from brain features). TMLE, a doubly-robust causal inference method, allowed an examination of the causal contribution of brain morphometry to binge drinking, over and above pure predictive value. This is rare in the field of neurodevelopment, with our recent review finding very little application of causal inference methods in the literature (16). Applying this novel machine learning/causal inference pipeline in two large and rich prospective datasets allowed for the comparison of findings across different stages of adolescent neurodevelopment.

### Limitations

It’s likely that some important confounders of the neurodevelopment–binge drinking relationship were missing from our causal inference analysis. That is, the assumption of conditional exchangeability required for valid causal inference may not hold. That said, given the estimates from these analyses were already small and non-significant, inclusion of additional confounders would be unlikely to alter findings in a meaningful way. However, we cannot rule out bias from violation of the other assumptions for valid causal inference: positivity (that for any given set of covariate values, it must be possible to be both a binge drinker or an abstainer), consistency (that there is only one ‘version’ of the value of a particular morphometric feature), and no interference (that bingeing outcome for a given individual isn’t affected by other adolescents’ drinking).

Finally, it should be noted that individuals included from NCANDA provided baseline measurements across a wide age bracket (12-17 years), rather than a single age/narrow span as in ABCD. This means that brain morphometry in NCANDA analyses captures a mixture of neurodevelopmental stages, limiting inference about the predictive utility/causal contribution of the brain at any discrete adolescent age.

### Future research

Identifying risk factors for binge drinking and other escalated alcohol consumption behaviours in adolescence remains crucial, especially given drinking likely compounds any underlying neuroanatomical vulnerabilities (41). Implementing more abstracted and sophisticated brain-based analyses such as network-based methods, as well as an incorporation of resting state functional connectivity predictors (40,42), may be a fruitful avenue for future work. It could also be informative to focus just on those adolescents with apparent neuroanatomical ‘vulnerability’ at baseline and identify any ‘resilience factors’ that predict which of them avoid graduation to harmful alcohol use.

As the ABCD cohort ages, and more individuals engage in harmful alcohol use behaviours, greater research opportunities will arise. There may also be value in normative modelling approaches, which involve the calculation of a ‘normative’ reference curve of neurodevelopment over time, and focusing on how individuals’ deviations from this curve may predict substance use outcomes.

## Conclusions

The present study did not find evidence that early brain structure is causally implicated in the development of binge drinking in adolescence. When considered alongside other recent findings, it may be that social and familial factors exert stronger influences on alcohol use outcomes at this particular developmental stage.

## Data Availability

Researchers can apply for access to the data here: https://nda.nih.gov/abcd/request-access

## ACKNOWLEDGEMENTS

This research was supported by grants from the NIH’s NIAAA (Award numbers: R01AA030575, MPI: Mewton/Squeglia; K24AA031052, PI: Squeglia). Generative AI (Claude) was used to assist in construction of Figure 2, with authors reviewing its output and taking full responsibility for the content.

## DISCLOSURES

All authors have nothing to declare.

## Notes

### Competing Interest Statement

The authors have declared no competing interest.

### Author Declarations

The ABCD protocol was approved by the centralised institutional review board (IRB) at the University of California, San Diego and by IRBs at each of the 21 sites. Similarly, for NCANDA, IRBs at each site approved the study. All data were de-identified prior to analysis.

## REFERENCES

1. Charrier L, Van Dorsselaer S, Canale N, Baska T, Kilibarda B, Comoretto RI, et al. (2024): A Focus on Adolescent Substance Use in Europe, Central Asia and Canada. Health Behaviour in School-Aged Children International Report from the 2021/2022 Survey. Volume 3. World Health Organization. Regional Office for Europe.

2. Miech RA, Johnston LD, Patrick ME, O’Malley PM (2025): National Survey Results on Drug Use, 1975–2024: Overview and detailed results for secondary school students. Monitoring the Future.

3. Lees B, Meredith LR, Kirkland AE, Bryant BE, Squeglia LM (2020): Effect of alcohol use on the adolescent brain and behavior. Pharmacol Biochem Behav 192: 172906.

4. Steinberg L, Albert D, Cauffman E, Banich M, Graham S, Woolard J (2008): Age differences in sensation seeking and impulsivity as indexed by behavior and self-report: evidence for a dual systems model. Dev Psychol 44: 1764.

5. Casey BJ, Jones RM, Somerville LH (2011): Braking and accelerating of the adolescent brain. Journal of research on adolescence 21: 21–33.

6. Lees B, Garcia AM, Debenham J, Kirkland AE, Bryant BE, Mewton L, Squeglia LM (2021): Promising vulnerability markers of substance use and misuse: A review of human neurobehavioral studies. Neuropharmacology 187: 108500.

7. Honarvar F, Arfaie S, Edalati H, Ghasroddashti A, Solgi A, Mashayekhi MS, et al. (2023): Neuroanatomical predictors of problematic alcohol consumption in adolescents: a systematic review of longitudinal studies. Alcohol and Alcoholism 58: 455–471.

8. Lopez-Vergara HI, Merrill JE, Janssen T, Jackson KM (2017): Social and individual-level predictors of alcohol use initiation and escalation: replicating and extending tests of differential effects. J Stud Alcohol Drugs 78: 452–457.

9. Wierenga LM, Langen M, Oranje B, Durston S (2014): Unique developmental trajectories of cortical thickness and surface area. Neuroimage 87: 120–126.

10. Squeglia LM, Ball TM, Jacobus J, Brumback T, McKenna BS, Nguyen-Louie TT, et al. (2017): Neural predictors of initiating alcohol use during adolescence. American journal of psychiatry 174: 172–185.

11. Rane RP, de Man EF, Kim J, Görgen K, Tschorn M, Rapp MA, et al. (2022): Structural differences in adolescent brains can predict alcohol misuse. Elife 11: e77545.

12. Ferariu A, Chang H, Kumar A, Sahl A, Gorka S, Wang L, et al. (2025): Integrating multilevel, multidomain and multimodal neuroimaging factors to predict early alcohol exposure trajectories using explainable AI. Dev Cogn Neurosci 101597.

13. Brown SA, Brumback TY, Tomlinson K, Cummins K, Thompson WK, Nagel BJ, et al. (2015): The National Consortium on Alcohol and NeuroDevelopment in Adolescence (NCANDA): a multisite study of adolescent development and substance use. J Stud Alcohol Drugs 76: 895–908.

14. Squeglia LM, Cservenka A (2017): Adolescence and drug use vulnerability: findings from neuroimaging. Curr Opin Behav Sci 13: 164–170.

15. Zhao Q, Pohl KM (2026): Leveraging Machine Learning to Advance Alcohol Research: Current Applications, Challenges, and Opportunities. Alcohol Res 46: 03.

16. Visontay R, Squeglia LM, Sunderland M, Devine EK, Byrne H, Mewton L (2024): Enhancing causal inference in population-based neuroimaging data in children and adolescents. Dev Cogn Neurosci 70: 101465.

17. Hagler Jr DJ, Hatton S, Cornejo MD, Makowski C, Fair DA, Dick AS, et al. (2019): Image processing and analysis methods for the Adolescent Brain Cognitive Development Study. Neuroimage 202: 116091.

18. Desikan RS, Ségonne F, Fischl B, Quinn BT, Dickerson BC, Blacker D, et al. (2006): An automated labeling system for subdividing the human cerebral cortex on MRI scans into gyral based regions of interest. Neuroimage 31: 968–980.

19. Hagler Jr DJ, Ahmadi ME, Kuperman J, Holland D, McDonald CR, Halgren E, Dale AM (2009): Automated white matter tractography using a probabilistic diffusion tensor atlas: Application to temporal lobe epilepsy. Hum Brain Mapp 30: 1535–1547.

20. Mori S, Wakana S, Van Zijl PCM, Nagae-Poetscher LM (2005): MRI Atlas of Human White Matter. Elsevier.

21. Fortin J-P, Cullen N, Sheline YI, Taylor WD, Aselcioglu I, Cook PA, et al. (2018): Harmonization of cortical thickness measurements across scanners and sites. Neuroimage 167: 104–120.

22. Grummitt L, Visontay R, Clare P, Slade T, Birrell L (2025): Multivariable machine learning prediction of risky alcohol use in contemporary youth. Addiction 120: 2404–2412.

23. Schuler MS, Rose S (2017): Targeted maximum likelihood estimation for causal inference in observational studies. Am J Epidemiol 185: 65–73.

24. Díaz I, Williams N, Hoffman KL, Schenck EJ (2023): Nonparametric causal effects based on longitudinal modified treatment policies. J Am Stat Assoc 118: 846–857.

25. Qiu S, Hubbard AE, Gutiérrez JP, Pimpale G, Juárez-Flores A, Ghosh R, et al. (2024): Estimating the effect of realistic improvements of metformin adherence on COVID-19 mortality using targeted machine learning. Glob Epidemiol 7: 100142.

26. Nath N, Beltrano W, Haynes L, Dewey D, Bray S (2023): Long-term effects of preterm birth on children’s brain structure: an analysis of the adolescent brain cognitive development (ABCD) Study. eNeuro 10.

27. Polley E, LeDell E, Kennedy C, Lendle S, van der Laan M (2019): Package ‘superlearner.’ CRAN.

28. Williams N, Díaz I (2023): lmtp: an R package for estimating the causal effects of modified treatment policies. Obs Stud 9: 103–122.

29. Mowinckel AM, Vidal-Piñeiro D (2020): Visualization of brain statistics with R packages ggseg and ggseg3d. Adv Methods Pract Psychol Sci 3: 466–483.

30. Byrne H, Visontay R, Devine EK, Squeglia LM, Mewton L (2026): ggAtlasTrack: ggplot2 Visualisation Tools for AtlasTrack White Matter Tracts [no. 0.1.3].

31. Moore A, Lewis B, Elton A, Squeglia LM, Nixon SJ (2024): An investigation of multimodal predictors of adolescent alcohol initiation. Drug Alcohol Depend 265: 112491.

32. Green R, Wolf BJ, Chen A, Kirkland AE, Ferguson PL, Browning BD, et al. (2024): Predictors of substance use initiation by early adolescence. American journal of psychiatry 181: 423–433.

33. Boer OD, Franken IHA, Muetzel RL, Cousijn J, El Marroun H (2024): Examining associations between brain morphology in late childhood and early alcohol or tobacco use initiation in adolescence: Findings from a large prospective cohort. Biol Psychol 192: 108859.

34. Haber SN (2016): Corticostriatal circuitry. Dialogues Clin Neurosci 18: 7–21.

35. Miller AP, Baranger DAA, Paul SE, Garavan H, Mackey S, Tapert SF, et al. (2024): Neuroanatomical variability and substance use initiation in late childhood and early adolescence. JAMA Netw Open 7: e2452027.

36. Morales AM, Jones SA, Harman G, Patching Bunch J, Nagel BJ (2020): Associations between nucleus accumbens structural connectivity, brain function, and initiation of binge drinking. Addiction biology 25: e12767.

37. O’Brien JW, Hill SY (2017): Neural predictors of substance use disorders in Young adulthood. Psychiatry Res Neuroimaging 268: 22–26.

38. Yin HH, Knowlton BJ (2006): The role of the basal ganglia in habit formation. Nat Rev Neurosci 7: 464–476.

39. Byrne H, Visontay R, Devine EK, Wade NE, Jacobus J, Squeglia LM, Mewton L (2026): Brain network features predating early alcohol initiation in adolescence. Transl Psychiatry.

40. Moreira JFG, Allgaier N, Johnson ME, Potter A, Garavan H, Fair DA (2026): Connectome-based predictive modeling of concurrent and longitudinal substance use vulnerability in adolescence. Dev Cogn Neurosci 79: 101689.

41. Baranger DAA, Paul SE, Hatoum AS, Bogdan R (2023): Alcohol use and grey matter structure: Disentangling predispositional and causal contributions in human studies. Addiction biology 28: e13327.

42. Kardan O, Weigard AS, Cope LM, Martz ME, Angstadt M, McCurry KL, et al. (2025): Functional brain connectivity predictors of prospective substance use initiation and their environmental correlates. Biol Psychiatry Cogn Neurosci Neuroimaging 10: 203–212.

