## Supplement for "Exploratory brain-wide analyses predicting binge drinking: A machine learning and targeted causal inference pipeline in two adolescent cohorts"

Visontay *et al.*

**Covariate derivation**

| COVARIATES | NOTES |
| --- | --- |
| **ABCD** | |
| Age | Age reported at baseline, standardised |
| Sex | Sex at birth reported at baseline (male/female) |
| Ethnicity | A five-category variable reported at baseline: Hispanic, White, Black, Asian, Other |
| Highest parental education level across caregivers | A five-level variable reported at baseline: Up to high school (No diploma), High school diploma/GED, Some college, Bachelor’s degree, Graduate school/Professional degree |
| Religious prohibitions on alcohol | This question asked the caregiver if their child’s religion had rules forbidding use of alcohol, with options being Yes, No, or Don’t know (variable combined data from baseline and first follow-up, owing to large number of missing responses at baseline) |
| Baseline alcohol sipping | Whether the child reported ever sipping alcohol at baseline |
| Intracranial volume | Total intracranial volume at baseline, standardised |
| **NCANDA** | |
| Age | Age reported at baseline, standardised |
| Sex | Sex reported at baseline (male/female) |
| Race | A six-category variable reported at baseline: Native American, Asian, Pacific Islander, Black, White, Other  For TMLE analyses, Native American and Pacific Islander were collapsed with the Other category due to low cell counts |
| Hispanic ethnicity | A binary variable as to whether the child is of Hispanic ethnicity, reported at baseline |
| Parental education | Eight options for highest parental education level of parent attending at baseline: Less than high school, High school diploma or equivalency (GED), Associate degree (junior college), Bachelor’s degree, Master’s degree, Doctorate, Professional, Other  For TMLE analyses, Less than high school and High school diploma or equivalency (GED) were collapsed into a single category, as were Master’s degree, Doctorate and Professional, both due to low cell counts |
| Intracranial volume | Total intracranial volume at baseline, standardised |

**ComBat procedure**

Covariates to be included in later TMLE analyses were also included in ComBat harmonisation to preserve signals of interest (not including intracranial volume). ComBat was performed in both ABCD and NCANDA, and separately for each imaging metric (volume, surface area, cortical thickness, FA, MD, RD).

**Machine learning details**

10-fold, cross-validation repeated 5 times was performed, with nested 10-fold cross-validation for hyperparameter tuning (see Supplement for which hyperparameters were tuned) and nested 5-fold cross-validation for non-negative least squares weights for combining base learners. Age, sex, and intracranial volume were ‘regressed out’ of all morphometric predictors in the outer loop of the procedure, such that the resulting predictors reflect variance in regional brain features independent of these confounders. To address potential issues related to class imbalance, outer and inner cross-validation folds were stratified by binge drinking outcome so that both training and test sets maintained equal class proportions. Additionally, within the hyperparameter-tuning step for each base learner, the Synthetic Minority Oversampling Technique (SMOTE) was applied to the training portion of each inner fold to further address class imbalance.

**Base algorithms and tuning parameters**

The following base algorithms and parameters were tuned:

| Parameter | Options |
| --- | --- |
| **RANDOM FORESTS** |  |
| Number of features sampled at each tree split | Square root of number of predictors, number of predictors divided by 3 |
| Minimum node size to split at | 5, 10 |
| **ELASTIC NET** |  |
| Alpha (balance between ridge and lasso regression) | 0, 0.2, 0.4, 0.6, 0.8, 1 |
| Lambda (strength of regularisation penalty) | 0.01, 0.014, 0.019, 0.023, 0.0228, 0.032, 0.037, 0.041, 0.046, 0.05 |
| **SUPPORT VECTOR MACHINE** |  |
| Cost parameter | 0.1, 1, 10 |
| Sigma | 0.01, 0.1, 1 |
| **XGBOOST** |  |
| Eta (learning rate) | 0.1, 0.3 |
| Total number of boosting iterations | 50, 100 |
| Maximum depth of individual trees | 3, 6 |

**Importance score calculation**

Different methods for importance score calculation were used across the four base learners. For Random Forests and Support Vector Machine, permutation testing was employed, which involves randomly permuting the values in one variable at a time, to see how much overall model performance decreases. For elastic net, which is most analogous to traditional regression models, each predictor is assigned a beta coefficient (those removed from the model due to low contribution have a coefficient of zero). These beta coefficients were used as importance scores. For XGBoost, gain-based metrics are generated during the training phase that reflect average improvement in loss function that can be attributed to any given predictor.

**TMLE details**

TMLE involves separate estimation of both an *exposure model* (i.e., modelling exposure based on covariate values) and an *outcome model* (modelling outcome based on covariate values). The exposure model estimates how likely each individual was to receive their actual exposure and is used to generate a ‘clever covariate’ (similar to an inverse probability weight). The *outcome model* is used to predict outcomes for each individual under different values of exposure. Finally, the clever covariate is used to make a small, targeted adjustment to the predictions generated by the outcome model, improving accuracy and reducing bias in the overall effect estimate.

Machine learning was used to estimate both the exposure and outcome models for TMLE, each using a SuperLearner ensemble with 5-fold cross-validation (base learners were a generalised linear model, elastic-net, mean, Random Forests, and XGboost, with the first two removed for NCANDA due to perfect separation issues).

All categorical covariates were converted to dummy variables prior to TMLE. Those with missing covariate data were removed from the analysis (except for the religious alcohol prohibitions variable in ABCD, for which the missing indicator was retained owing to large amounts of missing data). When generating estimates of uncertainty, the nested nature of the data within study sites was incorporated via cluster-robust standard errors in ABCD, and block-bootstrapped confidence intervals in NCANDA (necessitated by the smaller sample size).

**Code**

Scripts for the machine learning and TMLE analyses are located here: https://github.com/Lifespan-and-brain-health-team/Binge-drinking-machine-learning-and-causal-inference
